# Automated Image Transcription for Perinatal Blood Pressure Monitoring Using Mobile Health Technology

**DOI:** 10.1101/2023.06.16.23291435

**Authors:** Nasim Katebi, Whitney Bremer, Tony Nguyen, Daniel Phan, Jamila Jeff, Kirkland Armstrong, Paula Phabian-Millbrook, Marissa Platner, Kimberly Carroll, Banafsheh Shoai, Peter Rohloff, Sheree L. Boulet, Cheryl G. Franklin, Gari D. Clifford

**Affiliations:** Department of Biomedical Informatics, Emory University, Atlanta, GA, USA; Center for Indigeous Health Research, Wuqu’ Kawoq — Maya Health Alliance, Tecpán, Chimaltenango, Guatemala; Department of Gynecology and Obstetrics, Emory University, Atlanta, GA, USA; Department of Obstetrics and Gynecology, Morehouse School of Medicine, Atlanta, GA, USA; Division of Global Health Equity, Brigham and Women’s Hospital, Boston, MA, USA; Department of Biomedical Engineering, Georgia Institute of Technology, Atlanta, GA, USA

## Abstract

This paper introduces a novel approach to address the challenges associated with transferring blood pressure (BP) data from oscillometric devices used in self-measured BP monitoring systems. The primary objective of this study is to improve the accessibility and usability of BP data for monitoring and managing BP during pregnancy and postpartum, particularly in low-resource settings. To this end, we developed an automated image transcription technique to effectively transcribe readings from BP devices. The photos of the BP devices were captured as part of perinatal mobile health (mHealth) monitoring systems, conducted in four studies across two countries. The Guatemala Set 1 and Guatemala Set 2 datasets include the data captured by a cohort of 49 lay midwives from 1697 and 584 pregnant women carrying singletons in the second and third trimesters in rural Guatemala during routine screening. Additionally, we designed an mHealth system in Georgia for postpartum women to monitor and report their BP at home with 23 and 49 African American participants contributing to the Georgia I3 and Georgia IMPROVE projects, respectively. We developed a deep learning-based model which operates in two steps: LCD localization using the You Only Look Once (YOLO) object detection model and digit recognition using a convolutional neural network-based model capable of recognizing multiple digits. We applied color correction and thresholding techniques to minimize the impact of reflection and artifacts. Three experiments were conducted based on the devices used for training the digit recognition model. Overall, our results demonstrate that the device-specific model with transfer learning and the device independent model outperformed the device-specific model without transfer learning. The mean absolute error (MAE) of image transcription on held-out test datasets using the device-independent digit recognition were 1.1 and 1.1 mmHg for systolic and diastolic BP in the Georgia IMPROVE and 1 and 0.6 mmHg in Guatemala Set 2 datasets. The MAE, far below the FDA requirement of 5 mmHg, makes the proposed model suitable for general use when used with appropriate error devices.

## Introduction

Hypertensive disorders of pregnancy (HDP) are the most common medical complication encountered during pregnancy [1]. HDPs are related to a combination of maternal, placental and fetal factors and can lead to serious complications which can cause maternal and fetal morbidity and mortality [2]. The burden of these complications is disproportionately borne by women in low and middle-income countries (LMICs) and resource-constrained areas of high-income countries. For example, in Latin America, pregnancy vascular disorders are the leading cause of maternal mortality where up to 26% of maternal deaths are estimated to be related to preeclampsia [3, 4]. In the USA, during 2017–2019, the prevalence of HDP among delivery hospitalizations increased from 13.3% to 15.9% [5]. This trend is particularly concerning given the existing disparities in maternal health outcomes across different regions. For example, Georgia has among the most disparate maternal health outcomes in the US with significant disparities in maternal morbidity and mortality rates and access to quality care [6].

These disparities are driven by a combination of social, economics and systematic factors [7–9]. Moreover, both the US and LMICs exhibit geographic and neighborhood-level disparities in hypertension burden [10]. These disparities highlight the importance of addressing systemic healthcare issues related to health equity in monitoring HDPs. Early detection, effective management, and timely referral to specialized care are essential to improve hypertension outcomes in pregnancy and reduce preventable maternal and fetal morbidity and mortality. However, there are limitations in management and control of hypertension in pregnancy which includes delay in the decision to seek care, failure to identify signs of high risk pregnancies along with a delay in responding to the clinical symptoms [11]. Traditionally, BP monitoring during pregnancy and postpartum is done through periodic visits to the healthcare provider. However, this approach may not always be feasible or practical, particularly in low-resource settings or for women with limited access to healthcare, and leaves gaps in care. The use of mobile health technology for BP monitoring during pregnancy and postpartum has the potential to address some of the challenges and disparities, enabling early detection and management of hypertension.

Routine BP monitoring has been shown to be an effective tool for identifying individuals at risk. BP self-measurement is often utilized as part of telemonitoring process that can help overcome issues related to poor healthcare access, white coat effect, and provide more detailed insights into the BP lability. However, this approach is also prone to errors through incorrect usage, poor choice of device and transcription and transmission errors [12]. In particular, most BP monitors have not been evaluated for operational accuracy in HDP, and those that have, often do not have easy and free Bluetooth connectivity [13]. This presents a key problem for home-based BP monitoring in pregnancy and elaborates the need for efficient and reliable methods for transcribing, reading, and transmitting data from standard BP devices.

This study aimed to address the challenges of BP monitoring during pregnancy and postpartum in populations with high rates of HDP and limited access to healthcare through the development of a low-cost and accessible mobile health (mHealth) system with automatic AI-based transcription of BP from LCDs. The developed BP image transcription model was trained and validated using the data collected in two countries, including in perinatal monitoring study in Guatemala [14, 15] and postpartum BP monitoring studies in Georgia, USA. Fig 1 shows the overview of the developed model and datasets used in each phase of training/validation and testing the model.

**Fig 1.**
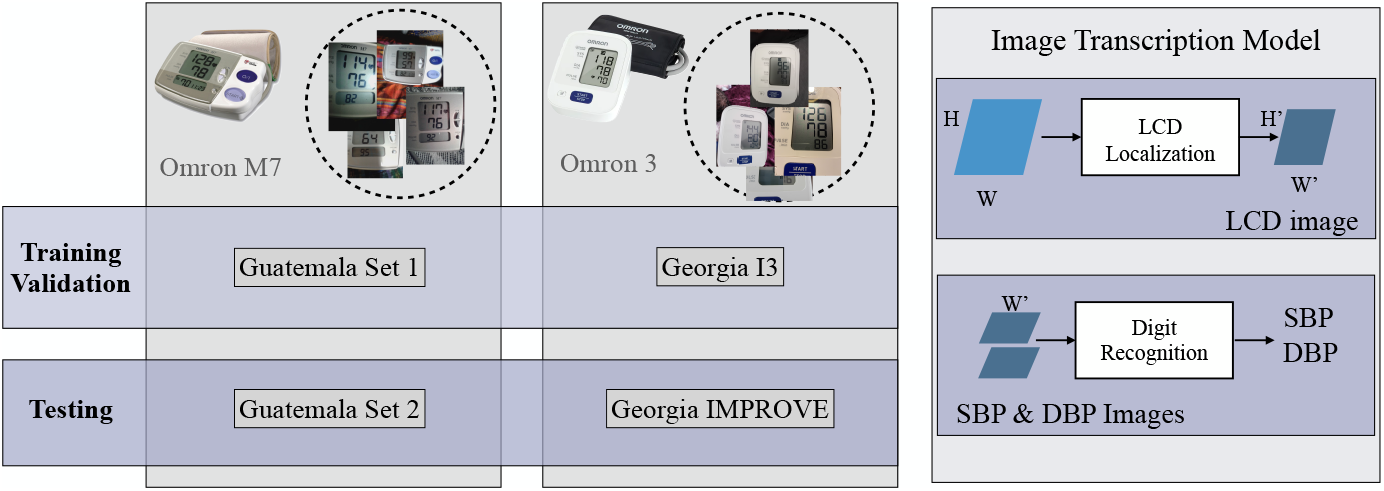
Overview of the model (right)and datasets utilized for model training, validation, and testing, including Guatemala Set 1, Guatemala Set 2, Georgia I3, and Georgia IMPROVE (left). The study employed an Omron M7 automated oscillometric BP monitor for the Guatemala datasets, and Omron 3 devices for the Georgia datasets. The BP image transcription model consists of two steps: 1) LCD Localization and 2) Digit recognition.

In the designed mHealth system the transmission of the BP measurement is based on using ubiquitous cell phone cameras. The developed deep-learning-based digit recognition model can automate transcribing the images. The proposed model includes LCD localization, pre-processing and classification of digits and has the potential to be applied in a wide range of applications where digit recognition in LCDs is needed, such as in glucose or weight measurement devices. By monitoring BP outside of clinical visits, and outside of office hours, we have the potential to capture BP in pregnancy and postpartum at critical, typically unmonitored, times. This mHealth solution is entirely scalable with no required specialized equipment and has the potential to improve maternal and fetal outcomes by enhancing access to accurate BP monitoring in low resource settings.

## Background

### mHealth system for BP monitoring

mHealth BP monitoring systems have demonstrated superior performance in comparison to traditional methods of BP monitoring, particularly in terms of convenience and management of hypertension [16, 17]. In these monitoring systems, once BP data has been measured, several methods allow users to record and transmit this data to clinicians. Core elements of number digitization are manual transcription on both paper and smartphone, Bluetooth or cellular data receivers and memory-card based and USB transfer. Each of these approaches has potential benefits and drawbacks, particularly in terms of risk of missing and inaccurate data. In the manual transcription, users may introduce errors during the transfer of data from the device display [18].

Furthermore, even trained clinical experts make significant errors when transcribing medical information [19]. Transferring the data using wireless BP devices is also prone to connectivity errors due to interference, variations in standards and various installed apps and services interfering with the connection. Memory card-based storage and USB transfer also introduce complications due to using cables. More importantly, given the implications of inaccurate BP measurement, validation of BP devices in hypertensive populations, especially perinatal populations, should be verified. The definitive work evaluating devices in hypertensive populations identified only a very small number of devices which are appropriate for preeclampsia, and none with wireless connectivity [13].

In earlier work, we showed that deep-learning-based digit recognition from photos of medical device displays can help accurately capture such data [20]. Automatic transcription of the images also facilitates home-based BP monitoring in populations with lower educational attainment and who are less likely to use mHealth tools, potentially due to challenges related to digital literacy.

### Digit Recognition

Digit recognition refers to the process of identifying handwritten or printed digits using machine learning algorithms. Optical Character Recognition (OCR) is one of the essential computer vision applications which involves converting images to editable and searchable digital documents. OCR technology has been in use since the 1980s [21–23]. Over the years, digit recognition algorithms have improved significantly and a variety of classifiers such as support vector machines, k-nearest neighbors, and neural networks have been used to recognize patterns in images and classify the numbers [24–26].

Among these algorithms, convolutional neural networks (CNN) have shown the best performance for digit recognition in various applications. In this work we developed an image-based OCR approach using CNN for accurate recognition of digits displayed on LCD screens.

## Data collection

In this study we trained and validated the BP image transcription model on four datasets which are described below.

### Guatemala Set 1

The Guatemala Perinatal mHealth Intervention study was conducted in rural areas of Guatemala to identify changes in outcomes of pregnant women due to the use of an Android mHealth app [14, 15]. At each visit, a traditional birth attendant recorded at least two maternal BP recordings using the Omron M7 (Omron Co., Kyoto, Japan) self-inflating device and captured the photo of the BP device using the developed mobile application (Fig 2). Visits were conducted in a mother’s home, where there might be poor lighting conditions. The user was trained to align the image using a mask that appears in the app for capturing the photo. Between January 2013 and July 2019, a total of 8,192 images were captured from 1,697 pregnant women carrying singletons between 6 weeks and 40 weeks gestational age. (This set of data is denoted ‘Guatemala Set 1’.) Before processing the images, the systolic BP, diastolic BP, and heart rate of each BP image were manually transcribed by three independent annotators. Annotators screened each of the images for readability as well as image quality labels. Readability was defined as the ability to clearly transcribe the full numerical values. A total of 7,205 images were annotated for the values of systolic BP (SBP), diastolic BP (DBP), heart rate (HR) along with a quality label. The distribution of BP readings is provided in Fig. 4. Segregation of these images based on their quality metric yielded 1,261 “Good Quality” images and 5944 poor quality images (inclusive of images with “Blur,” “Dark,” “Far,” “Contains Reflections,” and “Cropped” quality labels).

**Fig 2.**
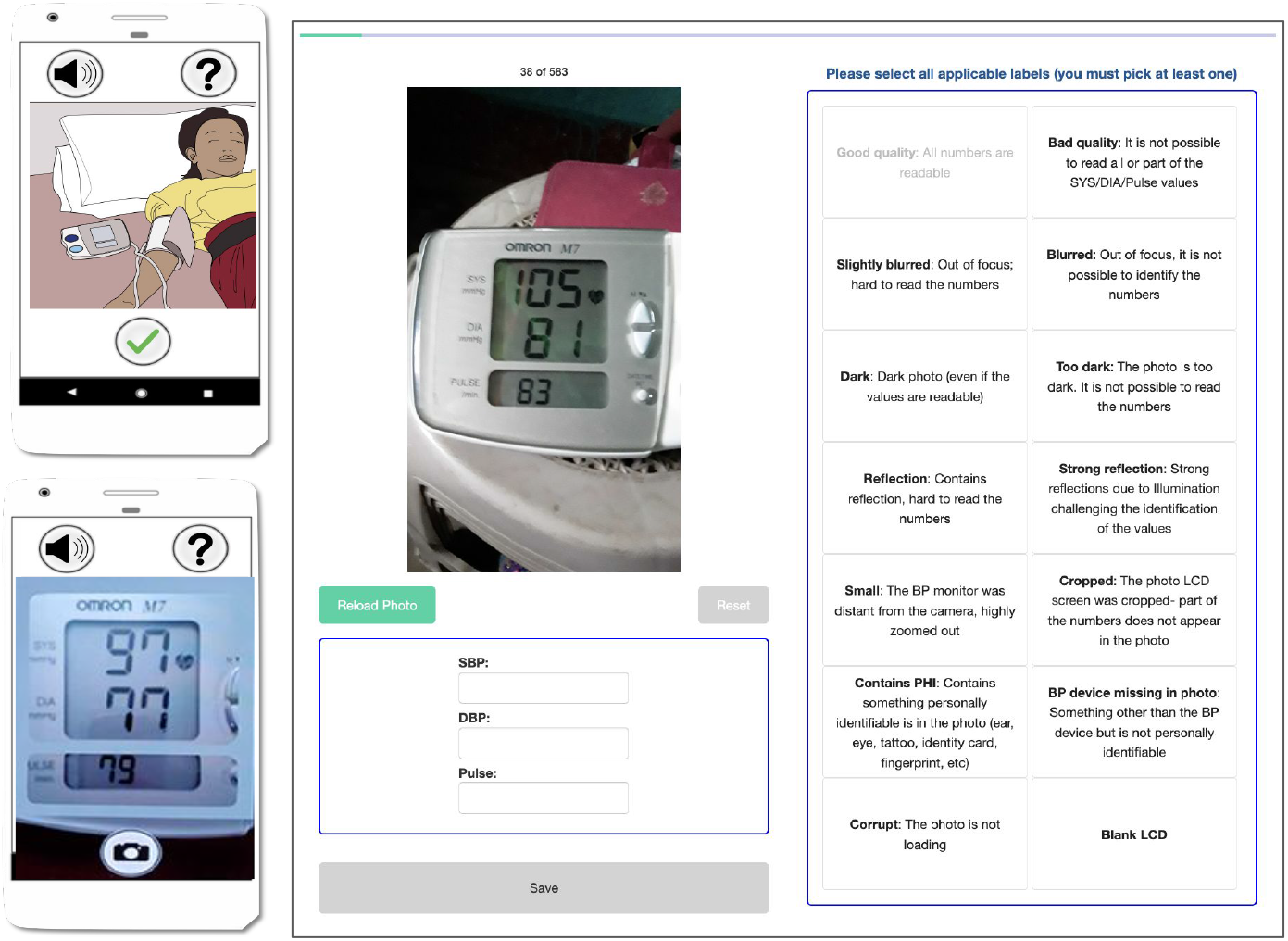
The designed mobile application and labeling web interface for transcribing and labeling the quality of the images in the Guatemala Perinatal study.

**Fig 3.**
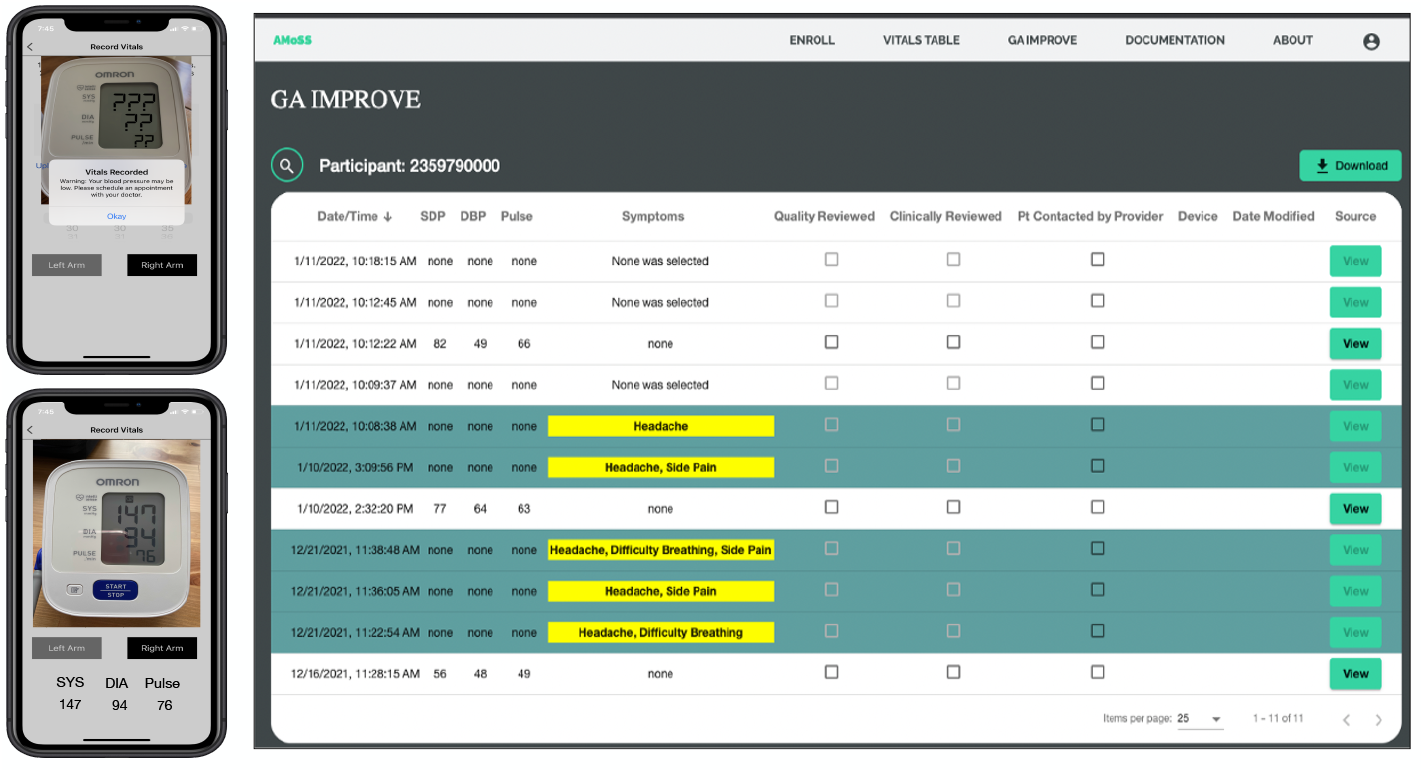
The backend dashboard and MoyoMom mobile app for capturing BP images and manual transcription in the Georgia I3 and Georgia IMPROVE studies for postpartum BP monitoring.

**Fig 4.**
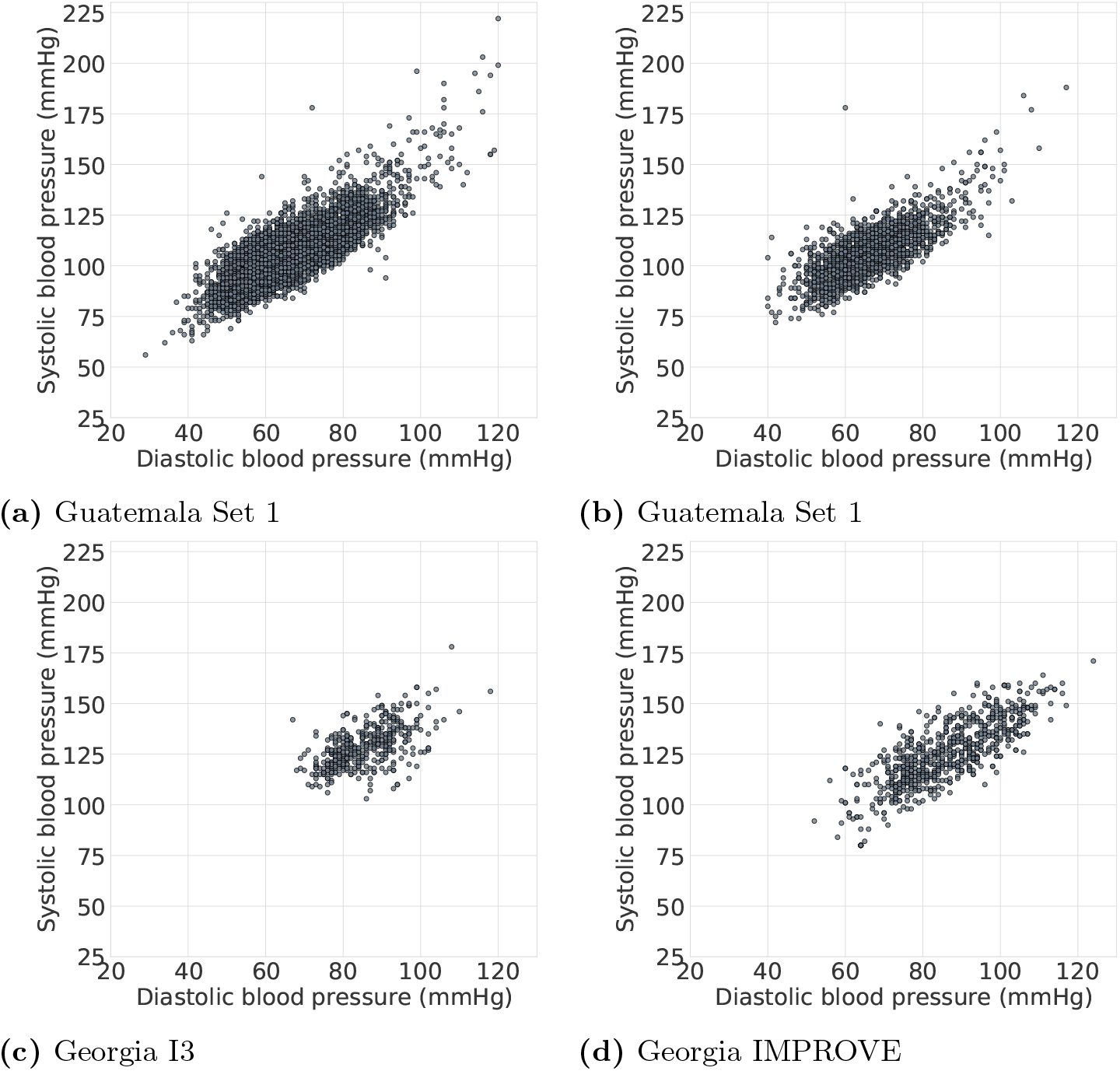
Distribution of BP data in the four datasets used in this study. The demographics of the individuals are given in table 1.

### Guatemala Set 2

Between August 2019 and October 2021, a total of 1934 blood pressure (BP) recordings were collected from 570 pregnant women by 28 midwives in Guatemala. (This set of data is denoted ‘Guatemala Set 2’.) The BP images were annotated by 10 independent annotators, with each image being labeled by three annotators. The labeling web interface was designed to collect the SBP, DBP and heart rate, with each of them being manually transcribed by the annotators. Additionally, the annotators labeled the quality of the images by choosing the defined quality labels. Fig 2 shows the examples of the app screens and the labeling interface.

### Georgia I3

The Georgia I3 was conducted in an urban setting in Georgia, US to study the feasibility of mHealth BP monitoring for the early detection of exacerbation of hypertension. Participants were postpartum women 18 years or older who delivered a liveborn infant at Grady Memorial Hospital in Atlanta, Georgia and were diagnosed with hypertensive disorders during pregnancy or at delivery. Consenting participants were given an Omron BP710N Series 3 upper arm BP monitor. Participants measured their BP twice daily for a 6-week period. They used the smartphone application (Moyo Mom) created by our team to capture the photo of the BP device and transcribe the numbers manually. A clinician had access to the participant data via a backend interface that serves as a case management portal (Fig 3). A study coordinator reviewed the collected data labeled the images as “study device”, “not study device” and “unidentified”. A study coordinator validated/corrected the numbers entered by participants using the designed clinical dashboard. The data consists of 475 BP images recorded by 23 participants. 427 images were captured from study devices, 36 other BP devices and 5 were unidentified images.

### Georgia IMPROVE

The Georgia IMPROVE study was designed to determine the association between cardiovascular complications during perinatal period, postpartum depression and symptoms of Covid-19. We recruited women 18 years or older during the late third trimester or early postpartum period in 11 sites in Georgia, including the Grady Memorial Hospital. The Moyo Mom app, used in the Georgia I3 study, was adapted for the IMPROVE study. The app, which is available on both iOS and Android platforms, is designed for participants to self-report metrics including but not limited to symptoms related to severe hypertension, effects of COVID-19 as well as personal experiences related to mental health, structural racism and discrimination. Participants are able to upload pictures of their BP readings which are then reviewed for accuracy by study coordinators and clinicians using the clinical dashboard. We have incorporated several alert features into the app design process in order to notify providers of potential poor outcomes, such as repeated high BP (*>*160/*>*110) readings, a reported symptom of severe hypertension and when a participant indicates self-harm ideation during the Mood Survey. Similar to Georgia I3, a study coordinator reviewed the collected data using the designed clinical dashboard. The collected data in this study includes 776 images from 48 participants where 720 images were captured from the study device, 6 other BP devices and 49 were unidentified images.

## Method

In this section the step-by-step approach to convert BP images into numerical format is described including LCD localization and the digit recognition methods.

### Automatic LCD Localization

Accurately localizing the LCD frames is essential for converting the images into a numerical format, but this can be challenging due to orientation and zooming effects, resulting in differences in the size and location of the frames. Object detection models, such as YOLO (You Only Look Once) [27], have demonstrated success in accurately localizing objects in images. To perform LCD localization using YOLO, the model was re-trained on a dataset of BP images for the task of LCD detection. During the training process, the model divides the image into a grid of cells and predicts the likelihood that an object, i.e., the LCD display, is present in each cell. The model also predicts the coordinates of the bounding box that surrounds the object, resulting in precise localization of the LCD display within the image.

### Digit Recognition

Our approach to transcribe the BP images is based on the recognition of sequence of digits in the LCD images. Specifically, we aim to learn a model of *P* (*S*|*X*) where *S* represents the output sequence and *X* represents the input image. To model *S*, we define it as *N* random variables *s*_1_, *s*_2_, …, *s*_*N*_ representing the elements of the sequence. In the task of BP transcription the maximum value of BP is a 3-digit number, therefore *N* is chosen to be 3 and each digit variable has 10 possible values. An additional “blank” character was incorporated for shorter sequences. In this work, the CNN-based model [20] was used to detect the sequences of digits. In this model a softmax classifier, receives extracted features from X by a convolutional neural network and returns the probability of each digit (Fig 5).

**Fig 5.**
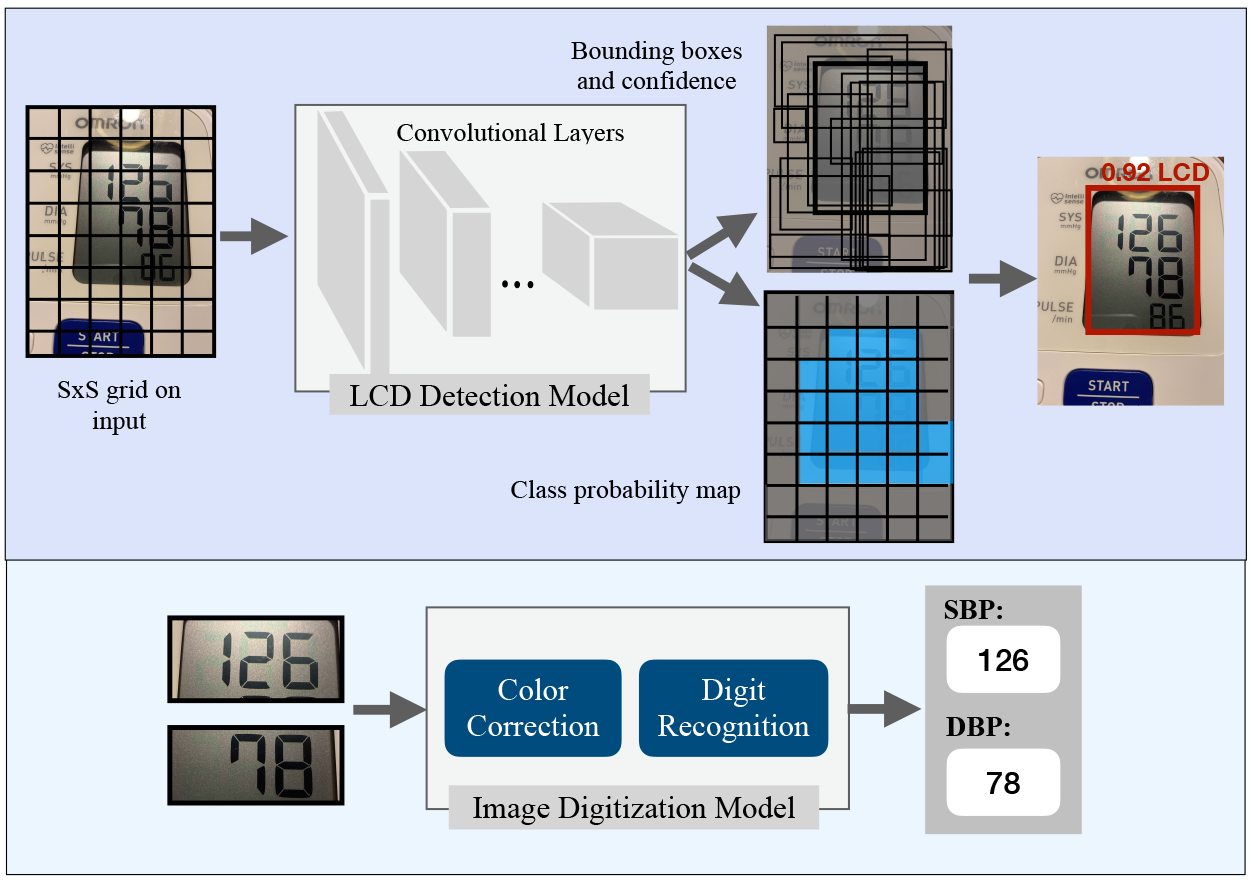
Overview of the BP image transcription. The LCD detection step uses the YOLO object detection model (top) and the CNN-based digit recognition model is for extracting numbers from SBP and DBP images (bottom).

### Experimental setup

The bounding box of the LCDs were annotated in 80 images from Guatemala Set 1 and Georgia I3 datasets (40 images per dataset) using the LabelImg toolbox [28]. Subsequently, the YOLO V5 model was re-trained using the labeled dataset for the task of LCD detection. Using the extracted LCDs, the SBP and DBP images were created and resized to a matrix size of 180 × 80. In the digit recognition model, a three-layer CNN architecture with 32, 64, 128 filters of dimension 5×5 was used and each layer was followed by batch normalization, ReLU activation, and maxpooling. The resulting feature vector from the CNN was then fed into three softmax classifiers. To optimize the model parameters, a sparse categorical cross entropy loss function and mini batch stochastic gradient descent were used.

### Model Evaluation

In the model evaluation, first, we compared the performance of the BP transcription using two different LCD localization methods: YOLO-based and contour-based [20] LCD detection.

Additionally, we investigated the effect of including LCD images extracted from two different BP devices (the Omron M7 and Omron 3) in the training phase. The performance of the digit recognition model was evaluated by defining three experiments and comparing the 5-fold cross validation results. As mentioned in the data collection section, the training and validation of the model were conducted using images captured from Omron M7 devices in the Guatemala Set 1 and Omron 3 BP devices in the Georgia I3 datasets. Following is the details of performed experiments:

- **Device-Specific:** Separate models were trained for each of the BP devices. Guatemala Set1 dataset was used to train the digit recognition model for the Omron M7 device and the Georgia I3 dataset was used for the device-specific model corresponding to the Omron 3 BP device. The five fold cross validation was used to assess the performance of the model.
- **Device-Specific with transfer learning:** In this experiment, training the model was based on using transfer learning. Specifically, we used a pre-trained model and fine-tuned the model. For example, in the digit recognition from the Omron M7 BP device, we used the model trained on images of Omron 3 and re-trained the model.
- **Device-Independent:** In this experiment, we merged the LCD images from both datasets and trained a single model. To evaluate the performance of the device-Independent model, we conducted a five fold cross validation on each dataset. In this evaluation, we included the images from the other BP device in the training set to assess the model’s robustness.

To evaluate the transcription performance, we used two evaluation metrics: classification accuracy and mean absolute error (MAE). Classification accuracy was defined as the percentage of correctly transcribed BP values out of the total number of samples. We also calculated the MAE between the automatic transcriptions generated by the model and validated BP values provided by the study coordinator or annotators depending on the dataset. Once the final model was trained, we utilized it to transcribe the images in the test datasets. The overview of the datasets used for training, validation and testing the model is presented in figure 1. To determine the statistical significance of the results, we performed the rank sum test to compare the manual and the automatic transcriptions. The null hypothesis was that there was no significant difference between the two methods, while the alternative hypothesis was that the automatic transcription was significantly different from the manual transcription.

## Results

We compared two different LCD localization methods and their impact on BP transcription accuracy. The YOLO-based method and the contour-based method were tested on the same set of data previously used in a study by Kulkarni et al. [20]. Our results show that the YOLO-based method outperformed the contour-based LCD localization method as shown in Table 2. In this experiment, the model was trained on 5020 single LCD images and tested on 1677 images. The results of BP transcription, showed that the YOLO-based method improved both the accuracy and MAE of transcribing SBP and DBP. This suggests that the YOLO-based method is more accurate in detecting LCDs in the images which leads to having better performance in BP transcription reducing the MAE of SBP and DBP detection to 1.04 and 0.91 mmHg, respectively. Fig 6 illustrates examples of the bounding boxes around the LCD screens detected using the YOLO-based method, along with the corresponding confidence scores generated by the model.

**Table 1.**
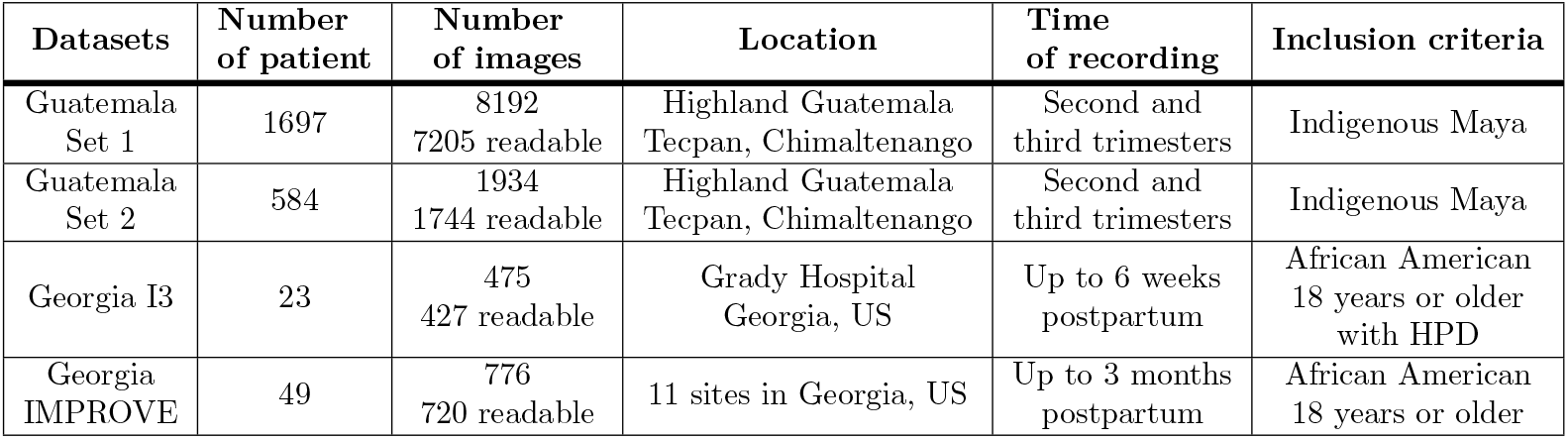
Datasets used for model training, validation, and testing.

| Datasets | Number of patient | Number of images | Location | Time of recording | Inclusion criteria |
| --- | --- | --- | --- | --- | --- |
| Guatemala Set 1 | 1697 | 8192<br>7205 readable | Highland Guatemala<br>Tecpan, Chimaltenango | Second and third trimesters | Indigenous Maya |
| Guatemala Set 2 | 584 | 1934<br>1744 readable | Highland Guatemala<br>Tecpan, Chimaltenango | Second and third trimesters | Indigenous Maya |
| Georgia I3 | 23 | 475<br>427 readable | Grady Hospital<br>Georgia, US | Up to 6 weeks postpartum | African American<br>18 years or older<br>with HPD |
| Georgia IMPROVE | 49 | 776<br>720 readable | 11 sites in Georgia, US | Up to 3 months postpartum | African American<br>18 years or older |

**Table 2.** Comparison of contour-based LCD localization and YOLO object detection method in performance of the BP image transcription in Guatemala perinatal data.

| LCD localization method: | Contour-based |  | YOLO-based |  |
| --- | --- | --- | --- | --- |
| Evaluation metrics | Acc | MAE | Acc | MAE |
| SBP | 90.7 | 3.19 | **93.7** | **1.04** |
| DBP | 91.1 | 0.94 | **96.6** | **0.91** |

**Fig 6.**
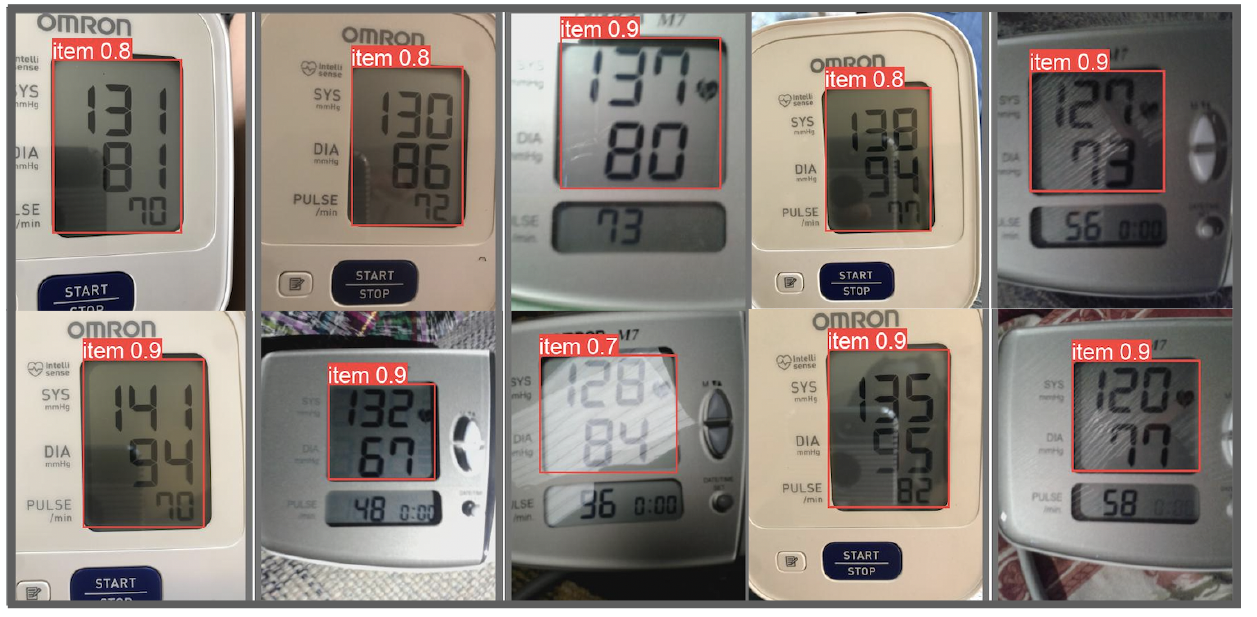
Examples of LCD detection results on images captured from Omron M7 and Omron 3 devices.

Our evaluation of the digit recognition model using three training strategies, as described in the “Model Evaluation” section, is summarized in Table 3 and Table 4. The results obtained through five-fold cross validation on the Guatemala Set 1 and Georgia I3 (Table 3), indicates that the best-performing models were achieved using transfer learning for each BP device and device independent model trained on images from both devices. The transfer learning model showed superior performance compared to the device independent model for the Georgia I3 dataset with an MAE of 0.6±0.4 and 1.1±1.6 mmHg for SBP and DBP, respectively. This might be attributed to the transfer learning model’s ability to compensate for the impact of low-quality images in the Guatemala Set 1.

**Table 3.**
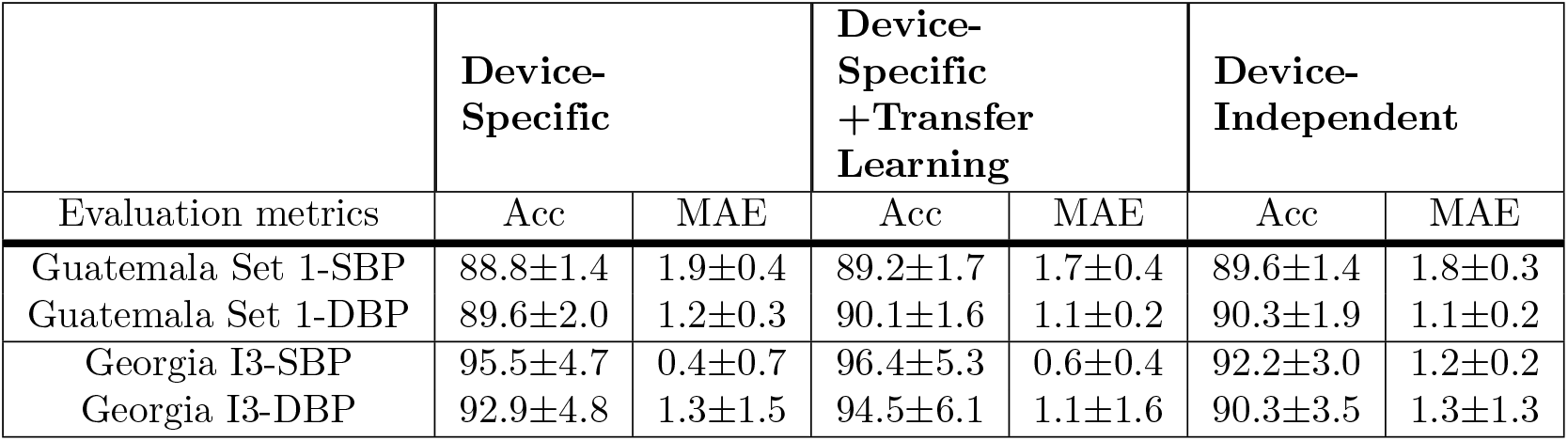
Five-fold cross validated results of the digit recognition model using three training strategies. Accuracy (Acc) is in percent and Mean Absolute Error (MAE) is in mmHg.

| Evaluation metrics | Device-Specific |  | Device-Specific +Transfer Learning |  | Device-Independent |  |
| --- | --- | --- | --- | --- | --- | --- |
|  | Acc | MAE | Acc | MAE | Acc | MAE |
| Guatemala Set 1-SBP | 88.8±1.4 | 1.9±0.4 | 89.2±1.7 | 1.7±0.4 | 89.6±1.4 | 1.8±0.3 |
| Guatemala Set 1-DBP | 89.6±2.0 | 1.2±0.3 | 90.1±1.6 | 1.1±0.2 | 90.3±1.9 | 1.1±0.2 |
| Georgia I3-SBP | 95.5±4.7 | 0.4±0.7 | 96.4±5.3 | 0.6±0.4 | 92.2±3.0 | 1.2±0.2 |
| Georgia I3-DBP | 92.9±4.8 | 1.3±1.5 | 94.5±6.1 | 1.1±1.6 | 90.3±3.5 | 1.3±1.3 |

**Table 4.** Testing the top model across folds on held-out test datasets. Accuracy (Acc) is in percent and Mean Absolute Error (MAE) is in mmHg.

| Evaluation metrics | Device-Specific |  | Device-Specific +Transfer Learning |  | Device-Independent |  | Average Human Transcription |  |
| --- | --- | --- | --- | --- | --- | --- | --- | --- |
|  | Acc | MAE | Acc | MAE | Acc | MAE | Acc | MAE |
| Guatemala Set 2-SBP | 93.3 | 1.0 | 93.7 | 1.3 | 93.6 | 1.0 | 93.1 | 4.1 |
| Guatemala Set 2-DBP | 93.1 | 0.7 | 93.8 | 0.5 | 94.0 | 0.6 | 92.6 | 2.7 |
| GA IMPROVE-SBP | 91.9 | 1.2 | 96.3 | 0.5 | 90.8 | 1.1 | 96.8 | 0.5 |
| GA IMPROVE-DBP | 86.2 | 3.5 | 92.7 | 1.8 | 92.0 | 1.1 | 96.9 | 0.3 |

In the next step, we assessed the performance of the optimized models on two held out test datasets, the Georgia IMPROVE and the Guatemala Set 2, as detailed in Table 4. We employed the MAE of SBP and DBP transcription as a metric to determine the top model across folds from the training/validation datasets. Overall, the device specific model with transfer learning and device independent models demonstrated the best performance. The result of the transfer learning approach was reported as MAE of 1.3 mmHg for SBP and 0.5 mmHg for DBP in the Guatemala Set 2 dataset and 0.5 mmHg, 1.8 mmHg for SBP and DBP in the Georgia IMPROVE dataset. And, using the device independent model the MAE was 1 mmHg for SBP and 0.6 mmHg for DBP in the Guatemala Set 2 dataset and 1.1 mmHg for both SBP and DBP in the Georgia IMPROVE dataset. These results demonstrate the capability of the developed model to accurately transcribe BP images. Our analysis indicates the superiority of incorporating images from two types of BP device, whether through the transfer learning approach or by merging the datasets in the training phase. The model’s performance on both held out test datasets underscores its effectiveness in capturing and generalizing important features, enabling it to provide precise SBP and DBP predictions.

We compared the manual and automatic transcriptions in the Georgia IMPROVE and the Guatemala Set 2 datasets. Detailed information regarding the manual transcription and validation of BP values for each dataset are provided in the “Data Collection” section. In the Georgia IMPROVE study, participants were instructed to input their BP values after capturing a photo of the device, and the study coordinator subsequently validated these transcriptions. For the Guatemala dataset, the data was transferred to our HIPAA compliant backend, where each image was labeled by three annotators. Table4 presents the MAE and accuracy metrics for an average human transcription. In the IMPROVE study, the analysis of manual transcription of SBP and DBP values resulted in an accuracy of 96.8% and 96.9%, respectively. The corresponding MAE values were 0.5 mmHg for SBP and 0.3 mmHg for DBP. In the Guatemala study, the accuracy of human transcription was lower than the IMPROVE study which might be due to lower quality of the images. The accuracy of the SBP and DBP annotations were 93.1% and 92.6% with MAE of 4.1 and 2.7 mmHg respectively. It should be noted that, during the processing of Guatemala Set 2, we removed images for which there was no agreement among the three annotators and the images with non-readable labels. Therefore, the reported results reflect the human transcription error for readable images with at least two consistent annotations. Considering all the images, we found that 91.9% and 91.3% of the images had consistent labels by all three annotators in annotating SBP and DBP values. In addition, we conducted a comparative analysis using a rank-sum test to assess the performance of the manual and the automatic transcriptions. In the evaluation of the Georgia IMPROVE dataset we found no statistically significant difference between the manual transcription by a single individual when compared to our automatic machine learning approach for both SBP (*p*-value=0.3) and DBP (*p*-value=0.1) transcriptions. Similar results were obtained for the Guatemala Set 2 dataset, with *p*-value of 0.4 and 0.5 for SBP and DBP respectively, as determined by the rank-sum test. These findings suggest that the developed automatic transcription method performs at a comparable level to manual transcription, demonstrating its potential as a reliable alternative.

## Discussion and Conclusion

While accuracies were generally greater than 90%, it is important to note that the error rates were generally very low, indicating that even when a transcription was incorrect, it was often in the last digit, and did not produce a clinically significant error. However, the error varied between datasets, which reflects the differences in both the lighting conditions (generally darker in less well-lit Guatemala homes) and the different devices. In particular, without retraining, the results exhibited lower performance on a different dataset. However, device-specific training with transfer learning and device-independent digit recognition models reduced the errors down to 1-2 mmHg, demonstrating that the errors are negligible (within the error bounds of the device itself). The device-specific approach is particularly useful when the type of device being used is known and can be taken into account during the transcription process. We note that our analysis demonstrates that the mean absolute error is far below the FDA requirement of 5 mmHg, which therefore makes the proposed model suitable for general use if the compound error with the chosen BP device remains within this limit. As such, we expect the continual updating of the model with more examples of a variety of BP models will eventually create a fully generalized model. In addition, we aim to enhance the model by adding an image quality assessment step which can provide real-time feedback to users to trigger recapture of data.

The integration of this technology into a clinical pathway for BP monitoring, recording and communication to healthcare professionals may enhance the management of hypertension and cardiovascular health. By automating the transcription of BP readings, this technology addresses critical challenges in capturing accurate data, particularly in low-literacy settings, and offers a range of transformative benefits.

Firstly, the automated transcription reduces the potential for human errors in the recording of BP measurements. By eliminating manual data entry, the technology can help to increase quality and consistency in the data captured. This, in turn, leads to more reliable diagnostic assessments and treatment decisions. Secondly, the developed model can enhance the efficiency and convenience of BP monitoring by simplifying the process of capturing and documenting BP readings. Moreover, the automated communication of BP data to healthcare professionals enables real-time monitoring and timely intervention. In conclusion, by mitigating errors, enhancing convenience, and enabling real-time communication, this innovative solution has the potential to significantly improve patient outcomes and strengthen the communication between patients and healthcare professionals.

## Data Availability

All data produced in the present study are available upon reasonable request to the authors

## Acknowledgments

Research reported in this publication was supported in part by the National Institutes of Health, the Fogarty International Center and the Eunice Kennedy Shriver National Institute of Child Health and Human Development (NICHD), grant number 1R21HD084114-01 (Mobile Health Intervention to Improve Perinatal Continuum of Care in Guatemala), NICHD grant number 1R01HD110480 (AI-driven low-cost ultrasound for automated quantification of hypertension, preeclampsia, and IUGR), the Imagine, Innovate and Impact (I3) Funds from the Emory School of Medicine and through the Georgia IMPROVE, funded by NIH National Center for Advancing Translational Sciences (NCATS) as an Administrative Supplement to the Georgia Clinical and Translational Alliance (UL1-TR002378). The content is solely the responsibility of the authors and does not necessarily represent the official views of the National Institutes of Health. We would also like to acknowledge the support of the Grady Health System, Atlanta, Georgia in conducting this research.

